# Bayesian hierarchical model for dose finding trial incorporating historical data

**DOI:** 10.1101/2022.03.12.22272175

**Authors:** Linxi Han, Qiqi Deng, Zhangyi He, Frank Fleischer, Feng Yu

**Affiliations:** School of Mathematics, University of Bristol, Bristol, United Kingdom; Biostatistics and Data Sciences, Boehringer Ingelheim Pharmaceuticals Inc., Ridgefield, Connecticut, USA; Department of Epidemiology and Biostatistics, School of Public Health, Imperial College London, London, UK; Biostatistics+Data Sciences Corp., Boehringer Ingelheim Pharma GmbH & Co. KG, Biberach, Germany

**Author notes:** Corresponding author. Email address (Feng Yu). 200 Technology Square Cambridge, MA 02139, United States. Cancer Research UK Beatson Institute, Glasgow G61 1BD, United Kingdom.

**Keywords:** Dose finding, Historical borrowing, Bayesian hierarchical framework, Between-trial heterogeneity

## Abstract

The Multiple Comparison Procedure and Modelling (MCPMod) approach has been shown to be a powerful statistical tool that can significantly improve the design and analysis of dose finding studies under model uncertainty. Due to its frequentist nature, however, it is difficult to incorporate into MCPMod information from historical trials on the same drug. Recently, a Bayesian version of MCPMod has been introduced by Fleischer et al. (2022) to resolve this issue, which is particularly tailored to the situation where there is information about the placebo dose group from historical trials. In practice, information may also be available on active dose groups from early phase trials, e.g., a dose escalation trial and a preceding small Proof of Concept trial with only a placebo and a high dose. To address this issue, we introduce a Bayesian hierarchical framework capable of incorporating historical information about both placebo and active dose groups with the flexibility of modelling both prognostic and predictive between-trial heterogeneity. Our method is particularly useful in the situation where the effect sizes of two trials are different. Our goal is to reduce the necessary sample size in the dose finding trial while maintaining its target power.

## 1. Introduction

A key component of successful pharmaceutical drug development is the accurate determination of an appropriate dose level in early phase clinical trials. The final selection of dose level in Phase II studies has a considerable impact on the success probability of confirmatory Phase III studies and consequently the entire drug development program. Owing to the limited amount of available data on the efficacy and safety of a specific compound and small sample sizes in early stages of clinical development, determining the right dose level is among the most challenging steps during drug development. As mentioned in Bretz et al. (2005), if the selected dose is too high, it can lead to safety problems as well as unacceptable adverse events in later studies, while a too-low dose leads to an increased likelihood that the drug fails to provide adequate clinical benefit, which impacts success probabilities of trials in later phases. Hence, when we evaluate the dose-response relationship, it is important to use a method capable of extracting the most information from available data. A method based on a statistical model that accurately reflects the clinical situation stands the best chance of yielding accurate estimates of the dose-response relationship.

Traditionally, dose-finding trials have been addressed through either a multiple comparison procedure (MCP) or a modelling (Mod) approach. Bretz et al. (2005) combined these two approaches to devise an improved method called Multiple Comparison Procedure and Modelling (MCPMod). The classic MCPMod assumes normally distributed data. Pinheiro et al. (2014) extended the approach to non-normal data. The main idea of MCPMod is to use a candidate set of parametric models to describe the unknown dose-response relationship, test these candidate models against a flat curve using MCP techniques and finally select a significant model to fit the data. This approach is reasonably robust against model misspecification since it allows the specification of multiple candidate models that can hopefully cover all plausible dose-response relationships. It is also flexible in the estimation of the optimal dose, which is not restricted to the doses under investigation. European Medicines Agency (EMA) (2014) and USA Food and Drug Administration (FDA) (2016) qualified MCPMod as an adequate and efficient method for dose finding trials.

MCPMod is based on the frequentist methodology. As a result, it can be difficult for a new study to incorporate information from historical trials on the same drug. Neuenschwander et al. (2010) pointed out that information from historical trials can be important to the design and analysis of a new trial. The incorporation of such historical information in the design and analysis of a new study can reduce the duration of the trial and/or the number of patients necessary to achieve the desired power, resulting in a reduction of overall cost (Schmidli et al., 2014). There has been increasing interest in incorporating such historical information into the design and analysis of clinical studies (Berry, 2006). Existing methods to incorporating historical information includes power priors (Ibrahim & Chen, 2000), meta-analytic analyses (Neuenschwander et al., 2010) and commensurate priors (Hobbs et al., 2012). When incorporating historical data, it is always desirable to design the framework in a way that is capable of modelling potential differences between historical and current data. In view of this, these approaches discount the information in the historical data to either a fixed degree or a degree determined dynamically according to between-trial heterogeneity.

There has also been work that uses a Bayesian framework to include historical data in the design and analysis of new trials. Fleischer et al. (2022) proposed an approach, known as Bayesian MCPMod (BMCPMod), that focuses on the situation where such historical information is coming only from the placebo dose group. BMCPMod essentially reproduces the results of the classic MCPMod for non-informative priors but allows for the incorporation of historical data if available, leading to an improvement in design efficiency. However, in practice, historical information of active dose groups may also be available from early phase trials, e.g., a dose-escalation trial and a preceding small Proof of Concept (PoC) trial with only a placebo and a high dose.

To address this problem, in this work we introduce a Bayesian hierarchical framework for the dose finding problem. It incorporates historical information from both placebo and active dose groups and accounts for both prognostic and predictive between-trial heterogeneity. One of the primary use cases of this model is for cases where the effect sizes of two trials are different. The goal is to reduce the necessary sample size in dose finding trials while maintaining its target power. The Bayesian hierarchical model we propose in this work performs the requisite analysis combining data from the two trials. This is referred to as the meta-analytic-combined approach by Schmidli et al. (2014).

The remainder of this paper is structured as follows. In Section 2, we introduce our Bayesian hierarchical model and review the Bayesian pooling model. In Section 3, we perform simulation studies to compare these two approaches in terms of precision, power and type I error. Finally, Section 4 provides a summary and discussion of results.

## 2. Methodology

### 2.1. Bayesian hierarchical model

We assume there are two trials, a current trial and a historical trial, with ***Y*** ^(*c*)^ and ***Y*** ^(*h*)^ denoting the data from each trial, respectively. We also assume there are a total of *K* + 1 unique dose groups 𝒟 = {*d*_0_, …, *d*_*K*_} in the two trials, including a placebo dose *d*_0_. Let *I*_*c*_ and *I*_*h*_ denote the indices of doses present in the current and historical trial, respectively. Let 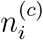 and 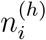 denote the number of patients from the current and or historical trial, respectively, who are given dose *d*_*i*_. For example, if *i* ∉ *I*_*c*_, then 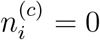. We assume the following statistical model for ***Y*** ^(*c*)^ and ***Y*** ^(*h*)^:

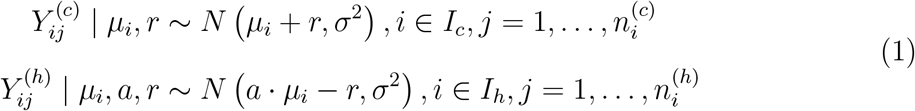

where *μ*_*i*_ = *μ*(*d*_*i*_) are the unknown mean effect of the treatment from the current trial at each dose level, *±r* denotes the deviation from the mean effect in each trial (as a result of heterogeneity of prognostic effects), and *a* is a scalar to allow for heterogeneity in the predictive effects between the two trials. The mean effect *μ*_*i*_ and the heterogeneity parameters *r* and *a* are assumed to be random. We will specify their prior distributions a bit later. We assume variance *σ*^2^ to be known and constant across all dose groups. The model can be easily extended to cases where the effect variance is not constant at different doses. Finally, we assume all 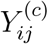 and 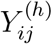 are conditionally independent given *μ*_*i*_, *r* and *a*.

For the mean effect ***μ*** = (*μ*_0_, …, *μ*_*K*_), we use an improper non-informative uniform prior *p*(*μ*_*i*_) ≡ 1 for all *i*. The heterogeneity of predictive effects (treatment effects) is represented by the fixed effect ratio *a* across different dose groups. The parameter *a* is assumed to be distributed as follows:

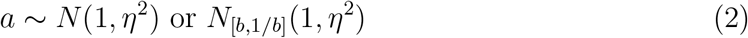

where *N*_[*b*,1*/b*]_ denotes the normal distribution with variance *η*^2^ truncated to the interval [*b*, 1*/b*] for some 0 *< b <* 1. The heterogeneity of prognostic effects between the two trials is expressed by the random effect *r* with the following prior

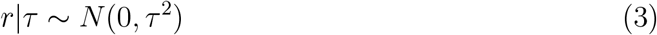

with between-trial standard deviation *τ*. The parameter *τ* controls the level of borrowing based on the similarity of prognostic effects between two trials. The prognostic effect under model (1) is 2*r* + (1 − *a*)*μ*_0_, where *μ*_0_ is the effect of the placebo dose.

We give some intuition about the choice of the hyperparameter *τ*. If *τ* is close to 0, then the assumption is that the prognostic heterogeneity is small between studies, i.e. there is a small difference in response of the placebo arm between studies. On the other hand, if *τ* is large, then the prognostic heterogeneity between the trials is high and the historical study data should have less relevance in the analysis of the data from the current trial. To account for the uncertainty about *τ*, we will use the half-normal prior distribution as its prior. This prior distribution should cover the range of typical values representing plausible (e.g., from small to large) between-trial heterogeneity (Spiegelhalter et al., 2004; Gelman, 2006; Rover et al., 2021). We will discuss the two types of heterogeneities in Sections 2.4 and 2.5. Given an observed outcome ***Y*** = (***Y*** ^(*c*)^, ***Y*** ^(*h*)^), the resulting posterior probability distribution for ***μ***, *a, r* and *τ* can be written as:

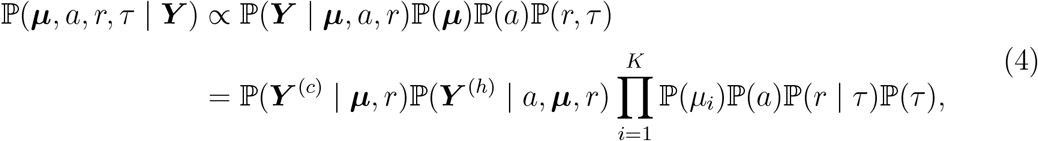

where

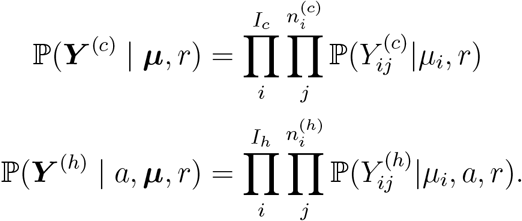

As the posterior probability distribution above is unavailable in a closed form, we perform MCMC sampling using the Metropolis-Hastings algorithm to obtain posterior samples of ***μ***, *a* and *r*. See Appendix A for details of this algorithm.

We recall that under the classic frequentist MCPMod, we would like to check for a non-flat relationship for the set of candidate models ℳ = {*M*_*m*_, *m* = 1, …, *M*}. We will define a Bayesian version of this test that corresponds to the null and alternative hypotheses, upon which we will assess type I error and power. Let ***c*** = {***c***_1_, …, ***c***_*m*_} be the contrast vectors for the candidate models ℳ and ***μ*** = (*μ*_1_, …, *μ*_*K*_) be the unknown mean effect vector of 𝒟. For model *m* and its corresponding contrast vector ***c***_*m*_, we consider the single model *M*_*m*_ to be significant at level *β* in our Bayesian sense if

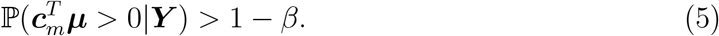

As in the case of the classic MCPMod, the maximum of these probabilities across all candidate models are considered in order to obtain our test decision. A significant doseresponse signal is established if

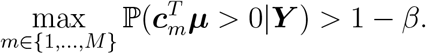

The value of *β* needs to be picked so that type I error of the overall procedure is equal to the significance level *α*, which is specified by the practitioner conducting the trial.

Under classic MCPMod, the probability on the left hand side of (5) can be computed numerically. This is, however, not feasible given the far more complicated model in our Bayesian setting. But we can approximate this probability using the ***μ***-samples generated using the MCMC procedure. If we have obtained a total of *N* samples ***μ***_1_, …, ***μ***_*N*_, we can simply compute 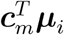 for each *i*. We count the number of samples that produces a positive 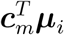 and divide it by *N*, which we take to be our estimate of

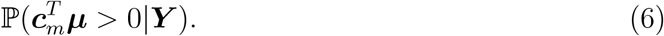

We summarise the procedure for checking the existence of non-flat relationship for the set of candidate models ℳ. This is the MCP part of the MCPMod. We fix a significance level *α*, then we perform the following:

Step 1: Define a set of candidate models to represent the underlying true dose-response shape, and derive the optimal contrast coefficients from each model based. This step mirrors corresponding steps in MCPMod.
Step 2: Generate datasets of historical and current trials assuming the null model (i.e. flat dose responses). Estimate the threshold *β* using steps leading to (6) so that the desired type I error rate *α* is achieved.
Step 3: With data ***Y***, test significance of each candidate model using (5) to assess whether a dose-response signal can be established. Proof of Concept (PoC) is established when at least one of the model tests is significant.

In this work, we focus on the MCP part of MCPMod. Given the ***μ***-samples we have generated for each candidate model in Step 3 above, we can perform the modelling part of the MCPMod using steps in the classic MCPMod. This is straightforward so we leave out the details.

### 2.2. Bayesian pooling model/ Bayesian model based on pooled data

An easy way to account for historical data is to simply pool the historical data with the current data, resulting in the pooled dataset ***Y*** ^(*p*)^. We end up with a special case of the Bayesian hierarchical model described in Section 2.1, with *r* = 0, ***Y*** ^(*p*)^ replacing ***Y*** ^(*c*)^, and the elimination of ***Y*** ^(*h*)^ and the parameter *a*. More specifically, 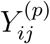 describes the variable of interest in patient *j* of dose group *i* and is assumed to be normally distributed

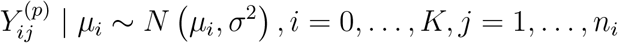

where *μ*_*i*_ are the unknown mean effect of the treatment and *σ*^2^ is assumed to be known.

We follow the same steps as described in Section 2.1 to perform the MCP step. If ***μ*** is given a non-informative prior, then the results obtained using our MCMC-based algorithm will converge to that of the classic MCPMod as the number of MCMC samples becomes large. We will compare our Bayesian hierarchical model with the Bayesian pooling model in Section 3.

### 2.3. Choice of contrast vectors

In classic MCPMod, contrast vectors are chosen to maximise the probability of rejecting the null hypothesis when a candidate model is correct. MCPMod is relatively robust against non-optimal choice of contrast vectors. More specifically, even when the candidate model set does not contain the true underlying dose-response model, the probability of detecting a non-flat dose-response shape is not greatly affected in the MCP step, as long as a dose-response model with a shape similar to that of the true shape of dose-response model is included in the candidate set.

In Pinheiro et al. (2014), MCPMod is extended to allow for an estimated 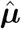, which can be estimated dose-response parameters using ANOVA or some other means. The estimates 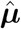 is assume to be distributed according to *N* (***μ, S***) where ***S*** denotes the variance/covariance of 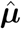. The matrix ***S*** also needs to be estimated, producing ***Ŝ***, which can be used to re-estimate the optimal contrast vectors. In our Bayesian setting, we use the posterior samples of ***μ*** to produce ***Ŝ***, which we use to re-estimate the optimal contrast vectors. Compared to the optimal contrast vectors calculated from the regular MCPMod, the re-estimated contrast vectors are data dependent and adjust the weight on difference dose groups automatically. For example, it will reduce the weight on doses where the posterior variance is large. As we will see in Section 3, this can improve the performance of our algorithm in certain cases.

We will use contrast vectors calculated according to the classic MCPMod as well as contrast vectors re-estimated using the posterior distribution in later simulation studies.

### 2.4. Heterogeneity of prognostic effects

In (1), heterogeneity of prognostic effects between different trials, which is defined to be independent of effects of treatment, is primarily accounted for by *r*. Generally speaking, when we design a clinical study, we typically have some prior information about how similar this study is to a historical study. This is usually assessed based on the similarity of key factors that could impact the response, for example, the target study populations, the regions of the site, and the formulation or delivery route of the intervention, etc. In our setting, as described in (3), we impose a normal prior with mean 0 and standard deviation *τ*. We impose a half-normal hyperprior on *τ*. Following Neuenschwander et al. (2010), we adopt 0.5*σ* as the standard deviation of the half-normal distribution. According to Friede et al. (2017), 0.5*σ* is sufficient to capture typical heterogeneity values seen in meta-analyses of heterogeneous studies, making it a sensible choice in many applications. The sensitivity analyses for different levels of heterogeneity prior will be discussed in Appendix E. Although we use a half-normal distribution here, a more flexible distribution could also be considered, e.g., half-T, half-Cauchy, Gamma or Inverse Gamma distributions (Spiegelhalter et al., 2004; Gelman, 2006; Polson & Scott, 2012).

### 2.5. Heterogeneity of predictive effects

In (1), the parameter *a* serves to model heterogeneity of predictive factors between different trials. It describes the difference in treatment effect sizes between studies. The maximum effect in the historical study is *a* times the one in the current study. Typically, predictive factors have a greater influence on the results of analysis than prognostic factors. As described in (2), we impose a normal prior distribution (truncated under some circumstances) on *a*, with mean 1 and between-trial heterogeneity standard deviation *η*. When *η* = 0 and *a* = 1, we have the same effect size between the historical and current trials, which means that there is no between-trial heterogeneity and therefore all data from two studies can be pooled for the analysis. The standard deviation *η* in the prior can be used to adjust the strength of prior information on *a*. We recommend plausible values of *a* to be inside the interval [1 − *η*, 1 + *η*], i.e. within one standard deviation of the mean 1.

We truncate the prior distribution for *a* to an interval [*b*, 1*/b*] for some 0 *< b <* 1 if ***Y*** ^(*c*)^ and ***Y*** ^(*h*)^ do not have exactly the same doses, i.e. there are doses that are present in one but the other trial. The reason for this is because in these cases, the posterior estimates for *a* are more unstable, thereby needing slightly more prior information to prevent *a* from becoming negative, which we deem as unrealistic. When we truncate the normal distribution, we typically pick *b* to be around 1/3, since it is unlikely that two trials differ in treatment effect size by a factor of more than 3. We do not recommend taking *b* to be very close to 0, as it leads to the simulated posterior distribution *a* to be concentrated at 0, resulting in non-convergence of the simulated posterior distribution of treatment effects of dose groups. We find the choice *b* = 1/3 gives robust results under our approach.

A log-normal distribution can also be used as the prior distribution of *a*. Since this distribution samples only positive values, its performance is similar to that of the truncated normal distribution.

## 3. Simulation studies

### 3.1. Simulation design

To evaluate the performance of our method, we run simulation studies. We take 𝒟_*c*_ = {0, 0.15, 0.5, 0.8, 1}, i.e. there are *K* = 4 active doses with a placebo dose in the current study. In the historic study, we consider two scenarios as described in Table 1a, with 4 and 5 doses that overlaps those in the current study. We standardise the doses and take the standard deviation to be 1. We take the true placebo response rate to be 0 and the maximal response to be 0.5. In what follows, we only consider equal allocation, i.e., each dose group contains 40 patients, 200 patients in total in the current study. We specify *M* = 6 different location-scale models in the candidate model set ℳ, as shown in Figure 1.

**Table 1:** Simulation scenarios.

| Scenario | Source | Dose set | Number of overlapping doses |
| --- | --- | --- | --- |
| 1 | Phase I dose escalation trial | $\mathcal{D}_h = \{0, 0.15, 0.5, 0.8, 1\}$ | 5 |
| 2 | Phase I dose escalation trial | $\mathcal{D}_h = \{0, 0.1, 0.5, 0.8, 1\}$ | 4 |

(a) Two scenarios of historical trial. Dose set of current trial $\mathcal{D}_c = \{0, 0.15, 0.5, 0.8, 1\}$ .
| Scenario | | $\Delta_c$ | $\Delta_h$ |
| --- | --- | --- | --- |
| A |  |  |  |
| Null hypothesis | $H_0^m : \mathbf{c}_m^T \boldsymbol{\mu} = 0$ | 0 | 0 |
| Alternative hypothesis | $H_1^m : \mathbf{c}_m^T \boldsymbol{\mu} > 0$ | 0.5 | 0.3 |
| B |  |  |  |
| Null hypothesis | $H_0^m : \mathbf{c}_m^T \boldsymbol{\mu} = 0$ | 0 | 0 |
| Alternative hypothesis | $H_1^m : \mathbf{c}_m^T \boldsymbol{\mu} > 0$ | 0.3 | 0.5 |
| C |  |  |  |
| Null hypothesis | $H_0^m : \mathbf{c}_m^{(c)T} \boldsymbol{\mu} = 0$ | 0 | 0.3 |
| Alternative hypothesis | $H_1^m : \mathbf{c}_m^{(c)T} \boldsymbol{\mu} > 0$ | 0.5 | 0.3 |

(b) Three hypothetical scenarios in simulation study. $\Delta_c$ = true effect size of current trial; $\Delta_h$ = true effect size of historical trial; $a = \Delta_h/\Delta_c$ ; $\mathbf{c}_m^{(c)}$ = contrast vector of current trial (elements of dose groups in historical trial only are zeros).

**Figure 1:**
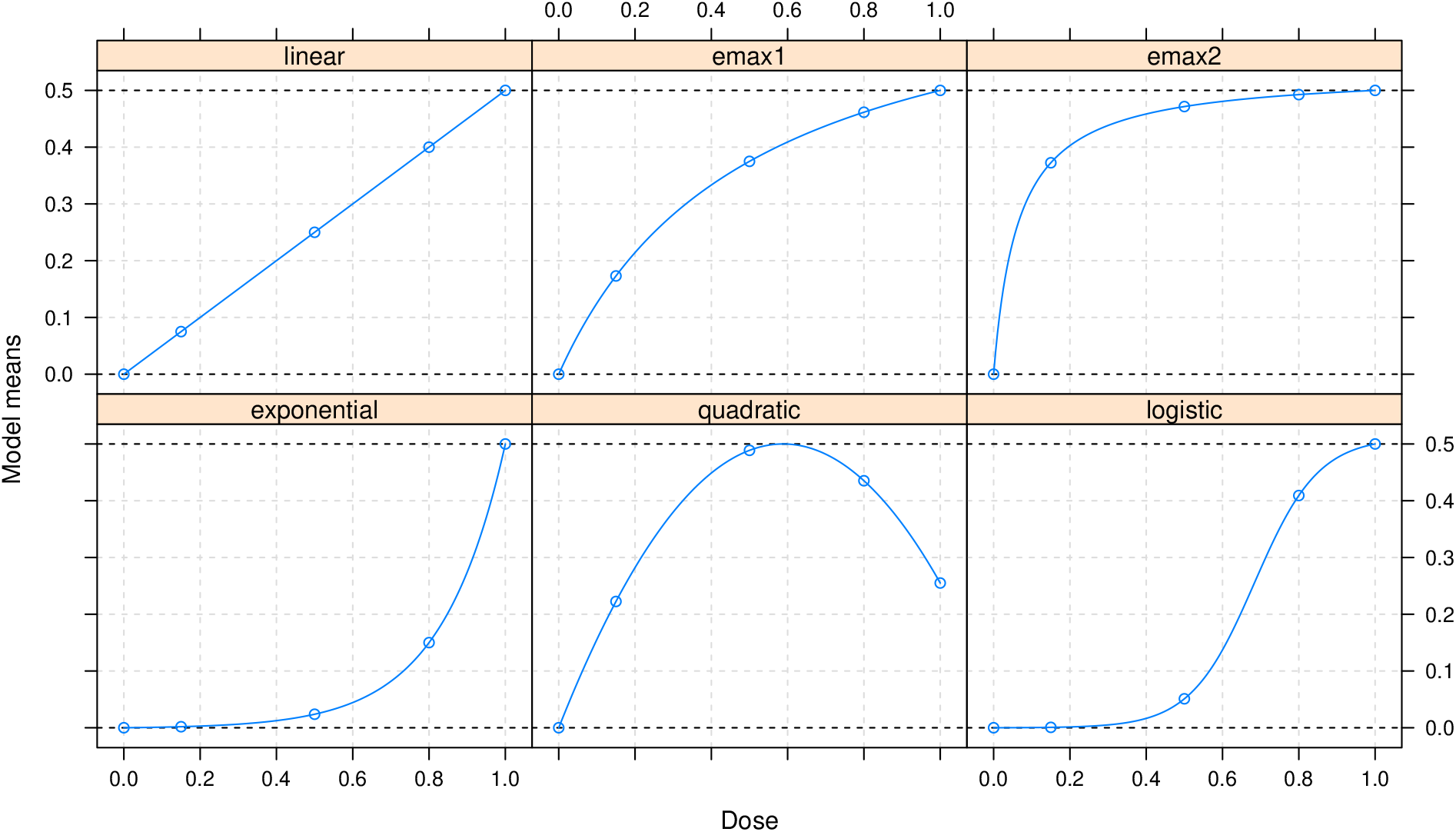
Visualisation of the candidate model set in the simulations.

Dose finding process in early stage usually contains a series of clinical trials for different purposes, including dose escalation trial, proof of concept (PoC) trial and dose-ranging trial. The Phase I dose escalation trial is usually the first time when a new drug is applied to humans. Such trial usually increases the doses of an investigational drug by cohorts, until an upper limit is reached, which is usually defined by the maximum tolerable dose. Sometimes it can also be based on a maximum feasible dose by formulation and delivery, or pharmacological active dose where the signal indicating the desired pharmacological effect is achieved. Although Phase I dose escalation trials are typically conducted in healthy volunteers and the efficacy data is not available, there are cases where the Phase I dose escalation trials need to be done in patients. Examples include investigating drugs for renal impaired patients where their PK profile will be different from healthy volunteers, or for drugs that may be toxic and cannot ethically be applied to healthy volunteers. After Phase I, a PoC needs to be established in early Phase II to make a Go/No-Go decision based on the efficacy performance. If the new drug demonstrates the efficacy, the concept is considered proven, leading to a Phase IIb dose-ranging trial. Therefore, a new dose-ranging trial can borrow historical information from a dose escalation trial or a PoC trial. In our simulation studies, we consider two scenarios: two dose escalation trials with different settings, as shown in Table 1a. We also consider an additional scenario (Phase IIa PoC trial) in Appendix D.

To evaluate the robustness of our method, we run three one-sided hypothesis tests in each scenario, which are summarised in Table 1b. Scenario A assumes the effect size of the historical trial is worse than the effect size of the current trial. The null hypothesis states that the treatment effect at each dose was the same as the placebo dose, and the alternative hypothesis states that at least one dose group has a positive treatment effect. Scenario B assumes that the treatment effect size of the historical trial exceeds the current trial. Definitions of the null and alternative hypotheses are the same as scenario A. In scenarios A and B, the results of the historical and current trials are basically homogeneous. The drug either worked in both trials or did not work in either.

In scenario C, however, we consider an extreme case of large discrepancies between the historical and the current trial. In scenarios A and B, the null and alternative hypotheses are built on the current and historical trials, but in scenario C, the null and alternative hypotheses are built on the current study only. More specifically, we take

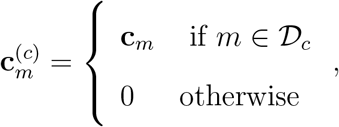

i.e. the test statistic does not take into account responses at doses not present in the current study. This scenario may arise from a situation where it is known that the drug has an effect in the historical trial and we investigate a new indication or population to find out whether the drug has an effect in the new trial. Differences in patient populations or other trial-specific circumstances can lead to large heterogeneity among historical trials and between the current trial and historical trials. In each scenario, we investigate four different levels of prognostic heterogeneity between the historical and current trials.

For each scenario, to assess the power to detect model *m* from the candidate model set ℳ, we simulate datasets assuming model *m* to be the true dose-response curve. The data is generated according to the treatment effect that is assumed under the alternative hypothesis for the computation of power, or according to the null hypothesis for type I error computation. The steps for estimating power and type I error of our algorithm are shown in Appendix B.

As described in Section 2.4, a half-normal prior with standard deviation 0.5 (i.e., *HN* (0.5)) is used for the between-study standard deviation of prognostic effects across scenarios A, B and C, and a truncated normal distribution with mean 1 and standard deviation 0.4 (i.e., *N*_[1/3,3]_(1, 0.4^2^)) is assigned for the effect ratio *a*. To compare our Bayesian hierarchical model (BHM), we also perform

1. a pooled Bayesian analysis that includes the data of all trials without accounting for between-trial heterogeneity,
2. an MCPMod analysis of only the current trial.

We use the Receiver Operating Characteristic (ROC) curve (Bradley, 1997; Fawcett, 2006) to evaluate and compare operating characteristics from different approaches. An ROC curve plots the True Positive Rate (TPR) (sensitivity), also known as power, against the False Positive Rate (FPR) (i.e., 1 – specificity), also known as type I error. In this paper, we plot ROC curves for FPR ∈ [0.025, 0.1], which is a reasonable type I error range for dose finding trials.

### 3.2. Simulation results

#### 3.2.1. Scenario 1

Here we consider a phase I dose escalation trial, where all dose groups in the historical trial are the same as those in the current trial, each with the same known variance and sample size. In Table 2a, we summarise powers achieved using various approaches at 5% type I error rate for the candidate model set illustrated in Figure 1. Amongst these approaches is MCPMod (full borrowing), where we simply pool data from both the historical and current studies to which we apply the classic MCPMod method. We show the corresponding ROC curves in Figure 4. For all dose groups, the type I error is calculated with a true flat dose-response curve with response 0, and the power is calculated with a true non-flat dose-response curve.

**Table 2:** Scenario 1.

| Approach | Linear | E<sub>max</sub>1 | E<sub>max</sub>2 | Exponential | Quadratic | Logistic |
| --- | --- | --- | --- | --- | --- | --- |
| MCPMod no borrowing | 0.7901 | 0.7916 | 0.7752 | 0.7638 | 0.6793 | 0.8681 |
| MCPMod full borrowing | 0.8759 | 0.8763 | 0.8650 | 0.8579 | 0.7839 | 0.9356 |
| BPM | 0.8710 | 0.8705 | 0.8651 | 0.8563 | 0.7744 | 0.9334 |
| BHM | 0.8964 | 0.8983 | 0.8951 | 0.8902 | 0.8185 | 0.9493 |

| Scenario | Model | $\vert r\vert = 0$ | $\vert r\vert = 0.1$ | $\vert r\vert = 0.2$ | $\vert r\vert = 0.3$ |
| --- | --- | --- | --- | --- | --- |
| Scenario 1A |  |  |  |  |  |
| Null hypothesis<br>($\Delta_c = \Delta_h = 0$) | | $r = 0$ | $r = 0.1$ | $r = 0.2$ | $r = 0.3$ |
|  | BHM | 0.0004 | 0.0027 | 0.0020 | 0.0016 |
|  | BPM | 0.0010 | -0.0006 | 0.0015 | 0.0023 |
| Alternative hypothesis<br>($\Delta_c = 0.5, \Delta_h = 0.3$) | | $r = 0$ | $r = 0.1$ | $r = 0.2$ | $r = 0.3$ |
|  | BHM | 0.4766 | 0.4782 | 0.4856 | 0.4892 |
|  | BPM | 0.4010 | 0.4019 | 0.4005 | 0.3971 |
| Scenario 1B |  |  |  |  |  |
| Null hypothesis<br>($\Delta_c = \Delta_h = 0$) | | $r = 0$ | $r = -0.1$ | $r = -0.2$ | $r = -0.3$ |
|  | BHM | 0.0011 | -0.0007 | 0.0009 | 0.0012 |
|  | BPM | 0.0001 | -0.0004 | 0.0016 | -0.0007 |
| Alternative hypothesis<br>($\Delta_c = 0.3, \Delta_h = 0.5$) | | $r = 0$ | $r = -0.1$ | $r = -0.2$ | $r = -0.3$ |
|  | BHM | 0.3828 | 0.3799 | 0.3752 | 0.3720 |
|  | BPM | 0.4002 | 0.4118 | 0.4011 | 0.3993 |
| Scenario 1C |  |  |  |  |  |
| Null hypothesis<br>($\Delta_c = 0, \Delta_h = 0.3$) | | $r = 0$ | $r = 0.1$ | $r = 0.2$ | $r = 0.3$ |
|  | BHM | 0.1157 | 0.1273 | 0.1201 | 0.1175 |
|  | BPM | 0.1509 | 0.1492 | 0.1458 | 0.1483 |
| Alternative hypothesis<br>($\Delta_c = 0.5, \Delta_h = 0.3$) | | $r = 0$ | $r = 0.1$ | $r = 0.2$ | $r = 0.3$ |
|  | BHM | 0.4718 | 0.4805 | 0.4863 | 0.4876 |
|  | BPM | 0.3992 | 0.4011 | 0.3998 | 0.4009 |

(b) Mean estimated treatment effect size under three simulation scenarios with four different levels of prognostic heterogeneity. $\Delta_c$ = effect size of current trial; $\Delta_h$ = effect size of historical trial. This table displays mean estimates of $\Delta_c$ .

We see from Table 2a and Figure 4 that MCPMod (no borrowing) yields the worst performance from the four approaches, which should be expected since borrowing from historical information can boost the power. MCPMod (full borrowing) and the Bayesian pooling model (BPM) have almost the same performance. In comparison, BHM yields better performance than all other approaches. This implies that modelling the between-trial heterogeneity can further improve the performance, i.e., BPM simply combines the historical and current trials into a single trial, but BHM discounts the historical information according to the between-trial heterogeneity. Note that here we use the optimal contrast vector derived from MCPMod rather than the re-estimated contrast vector, but these two contrasts are shown to reveal similar behaviours.

In the following, we will only focus on the analysis of the linear model. The other five models are expected to exhibit similar behaviour. The main purpose of the simulation study is to identify in each scenario which methods have good power. We generate data with four different levels of heterogeneity in prognostic factor effects, *r* = 0, 0.1, 0.2, 0.3, respectively. For each scenario, we estimate the treatment effect size of the current study, which is the mean of 10,000 posterior means of *μ*_5_. This is shown in Table 2b.

Figure 2 compares the ROC curve of BHM and BPM as a function of *FPR* ∈ [0.025, 0.1] for four cases with different heterogeneities of prognostic factor under scenario 1A, 1B and 1C. We see that the ROC curves of BPM under scenarios 1A and 1B look similar to each other. This is because the treatment effect size of the pooled trial is the average effect size of two trials, hence BPM is mainly influenced by the average treatment effect of two trials. For BPM, since the prognostic factor effect *r* has opposite signs in the historical and current trials, different levels of heterogeneity in prognostic factor effects have no influence on the results, so that similar patterns of results are observed across four different values of *r*. Table 2b shows that BHM produces more accurate estimates of the effect size as the value of *r* increases. This is probably due to the prior *HN* (0.5) we use for the prognostic between-trial heterogeneity standard deviation representing a large prognostics heterogeneity. Even though the bias becomes smaller with the increase of the value of *r*, the ROC curves of BHM look similar to each other for all four different values of *r*, as can be seen from Figure 2. The sensitivity analysis of the results for prognostic heterogeneity will be discussed in Appendix E.

**Figure 2:**
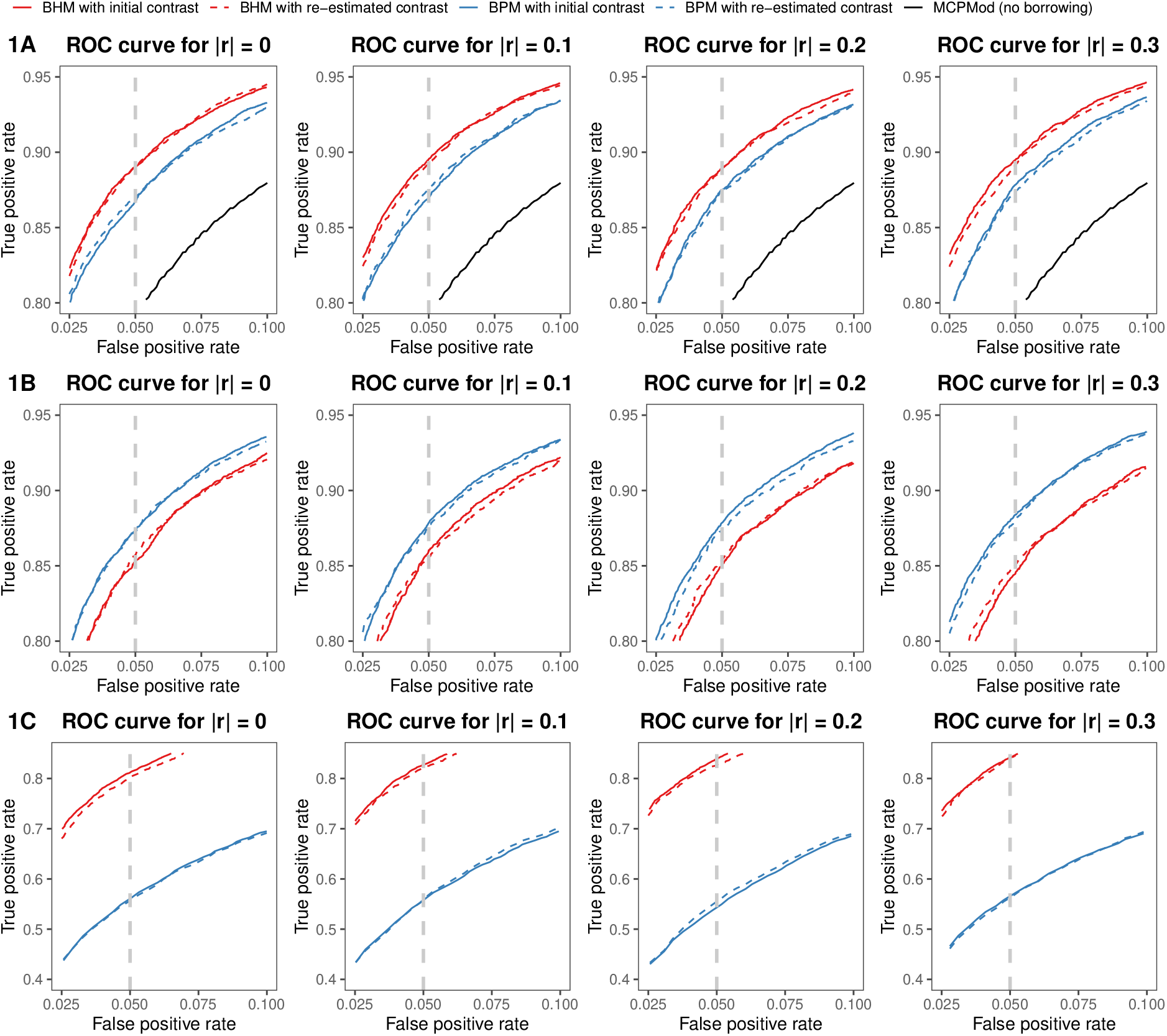
The ROC curves of MCPMod, Bayesian pooling model (BPM) and Bayesian hierarchical model(BHM) with four different levels of prognostic heterogeneity for scenario 1. The dashed grey line denotes 5% false positive rate, i.e., 5% type I error rate.

Under scenarios 1A and 1C, the ROC curve of BHM is always above the ROC curve of BPM, but under scenario 1B, BPM performs better than BHM. We surmise this is due to the fact that under scenario 1B where the effect size of the historical trial is higher than the current trial, BHM largely discounts the impact of historical data on the basis of observed divergence when incorporating historical information, leading to reduced power values. Even though BHM’s ROC curve is below that of BPM under scenario 1B, the estimates of the effect size under BHM is more accurate.

With BPM, when the treatment effect size in the new trial is worse than that in the historical trial, the borrowing of historical information leads to overestimates of treatment effects across the active dose groups. Thus it leads to higher power but that comes at the cost of type I error inflation. Generally speaking, we would prefer to use BHM to perform dynamic borrowing where the amount of historical data borrowed is related to the agreement between the current and historical trial.

When historical treatment effect size differs from the current trial, BPM may not reflect the true treatment effect size. On the other hand, the posterior mean estimates of the true effect size using BHM has smaller bias. Compared to BPM, BHM is a better statistical analysis method that can reasonably boost statistical power while controlling type I error.

#### 3.2.2. Scenario 2

Here we investigate a scenario where there is at least one non-overlapping dose between the historical and the current trial. In scenario 2, the current trial has a dose of 0.15 but not 0.1, and the historical trial has a dose of 0.1 but not 0.15. The existence of non-overlapping doses has an influence on both BPM and BHM.

Table 3 displays the estimated treatment effects of dose groups 0.1 and 0.15 under scenarios A, B and C. Apparently BHM produces more accurate estimates for these two dose groups, i.e. with smaller bias, than BPM. For both BHM and BMP, larger magnitudes in the level of prognostic heterogeneity *r* produces larger biases in the estimates.

**Table 3:** Estimated treatment effect of two non-overlapping doses under three simulation scenarios with four different levels of prognostic heterogeneity. This table displays mean estimates of *μ*_2_ or *μ*_3_. BHM = Bayesian hierarchical model; BPM = Bayesian pooling model.

| Scenario | Model | $\vert r\vert = 0$ | $\vert r\vert = 0.1$ | $\vert r\vert = 0.2$ | $\vert r\vert = 0.3$ |
| --- | --- | --- | --- | --- | --- |
| Scenario 2A |  |  |  |  |  |
| Null hypothesis | | $r = 0$ | $r = 0.1$ | $r = 0.2$ | $r = 0.3$ |
| 0.10 ($\mu_2 = 0$) | BHM | 0.0058 | -0.0206 | -0.0247 | -0.0269 |
|  | BPM | 0.0035 | -0.1016 | -0.2006 | -0.3015 |
| 0.10 ($\mu_3 = 0$) | BHM | 0.0005 | 0.0168 | 0.0139 | 0.0121 |
|  | BPM | 0.0022 | 0.1004 | 0.1976 | 0.3032 |
| Alternative hypothesis | | $r = 0$ | $r = 0.1$ | $r = 0.2$ | $r = 0.3$ |
| 0.10 ($\mu_2 = 0.030$) | BHM | 0.0220 | -0.0264 | -0.0328 | -0.0375 |
|  | BPM | 0.0283 | -0.0714 | -0.1705 | -0.2681 |
| 0.15 ($\mu_3 = 0.075$) | BHM | 0.0820 | 0.0980 | 0.1019 | 0.1012 |
|  | BPM | 0.0807 | 0.1735 | 0.2725 | 0.3726 |
| Scenario 2B |  |  |  |  |  |
| Null hypothesis | | $r = 0$ | $r = -0.1$ | $r = -0.2$ | $r = -0.3$ |
| 0.10 ($\mu_2 = 0$) | BHM | 0.0403 | 0.0255 | 0.0241 | 0.0258 |
|  | BPM | 0.0043 | 0.1021 | 0.2147 | 0.2975 |
| 0.15 ($\mu_3 = 0$) | BHM | 0.0004 | -0.0174 | -0.0159 | -0.0095 |
|  | BPM | 0.0046 | -0.1127 | -0.1965 | -0.3110 |
| Alternative hypothesis | | $r = 0$ | $r = -0.1$ | $r = -0.2$ | $r = -0.3$ |
| 0.10 ($\mu_2 = 0.050$) | BHM | 0.0519 | 0.0661 | 0.0683 | 0.0722 |
|  | BPM | 0.0507 | 0.1528 | 0.2439 | 0.3517 |
| 0.15 ($\mu_3 = 0.045$) | BHM | 0.0407 | -0.0294 | -0.0650 | -0.0803 |
|  | BPM | 0.0450 | -0.0547 | -0.1525 | -0.2526 |
| Scenario 2C |  |  |  |  |  |
| Null hypothesis | | $r = 0$ | $r = 0.1$ | $r = 0.2$ | $r = 0.3$ |
| 0.10 ($\mu_2 = 0.030$) | BHM | 0.0292 | -0.0310 | -0.0402 | -0.0414 |
|  | BPM | 0.0276 | -0.1288 | -0.2276 | -0.3318 |
| 0.15 ($\mu_3 = 0$) | BHM | 0.0010 | 0.0307 | 0.0526 | 0.0757 |
|  | BPM | -0.0003 | 0.0963 | 0.2006 | 0.2988 |
| Alternative hypothesis | | $r = 0$ | $r = 0.1$ | $r = 0.2$ | $r = 0.3$ |
| 0.10 ($\mu_2 = 0.030$) | BHM | 0.0220 | -0.0264 | -0.0328 | -0.0375 |
|  | BPM | 0.0283 | -0.0714 | -0.1705 | -0.2681 |
| 0.15 ($\mu_3 = 0.075$) | BHM | 0.0820 | 0.0980 | 0.1019 | 0.1012 |
|  | BPM | 0.0807 | 0.1735 | 0.2725 | 0.3726 |

Figure 3 displays ROC curves of both BHM and BPM, using both the initial contrast of MCPMod as well as re-estimated contrasts. Unlike in Scenario 1, we see that both BHM and BPM performs better if we use re-estimated contrast. We will focus on performance using re-estimated contrasts in the subsequent discussion. If there is no prognostic heterogeneity (i.e., *r* = 0), we find BHM to have mixed performance compared to that of BPM, sometimes better, sometimes worse. Indeed, when *r* = 0, the pooling model is the correct model, but this is not the case if *r* ≠ 0. Therefore we see the advantage of BHM over BPM tends to increase as the level of prognostic heterogeneity increases. This is caused by a decrease in the power of BPM as *r* increases, while the power of BHM holds roughly steady. When type I error is controlled at 5%, the power of BPM went from 0.8833 for *r* = 0 to 0.8614 for *r* = 0.3.

**Figure 3:**
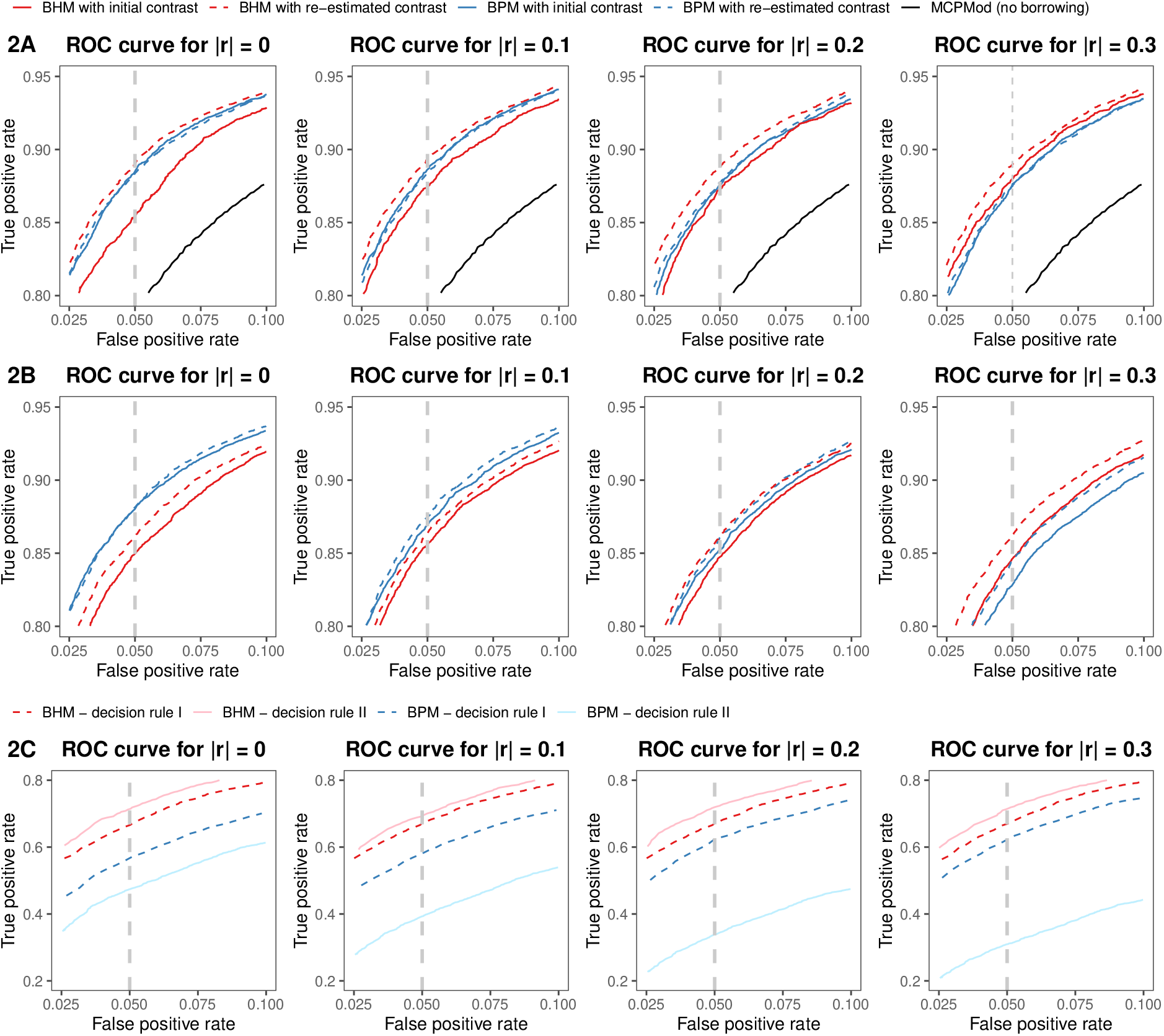
The ROC curves of MCPMod, Bayesian pooling model (BPM) and Bayesian hierarchical model (BHM) with four different levels of prognostic heterogeneity for scenario 2.

As mentioned earlier, we use re-estimated contrast vector in this scenario, not the initial contrast vector derived from MCPMod which would lead to some power loss. Figure 5 displays boxplots of the estimated treatment effects of six dose groups for BPM and BHM. The estimates for the second and third doses, both of which only exists in one of the two trials, has larger variability. Results from scenarios 2A, 2B and 2C show similar effect. Therefore, it is beneficial to apply a re-estimation step based on the covariance matrix **Ŝ** after each MCMC simulation to derive a re-estimated contrast vector.

**Figure 4:**
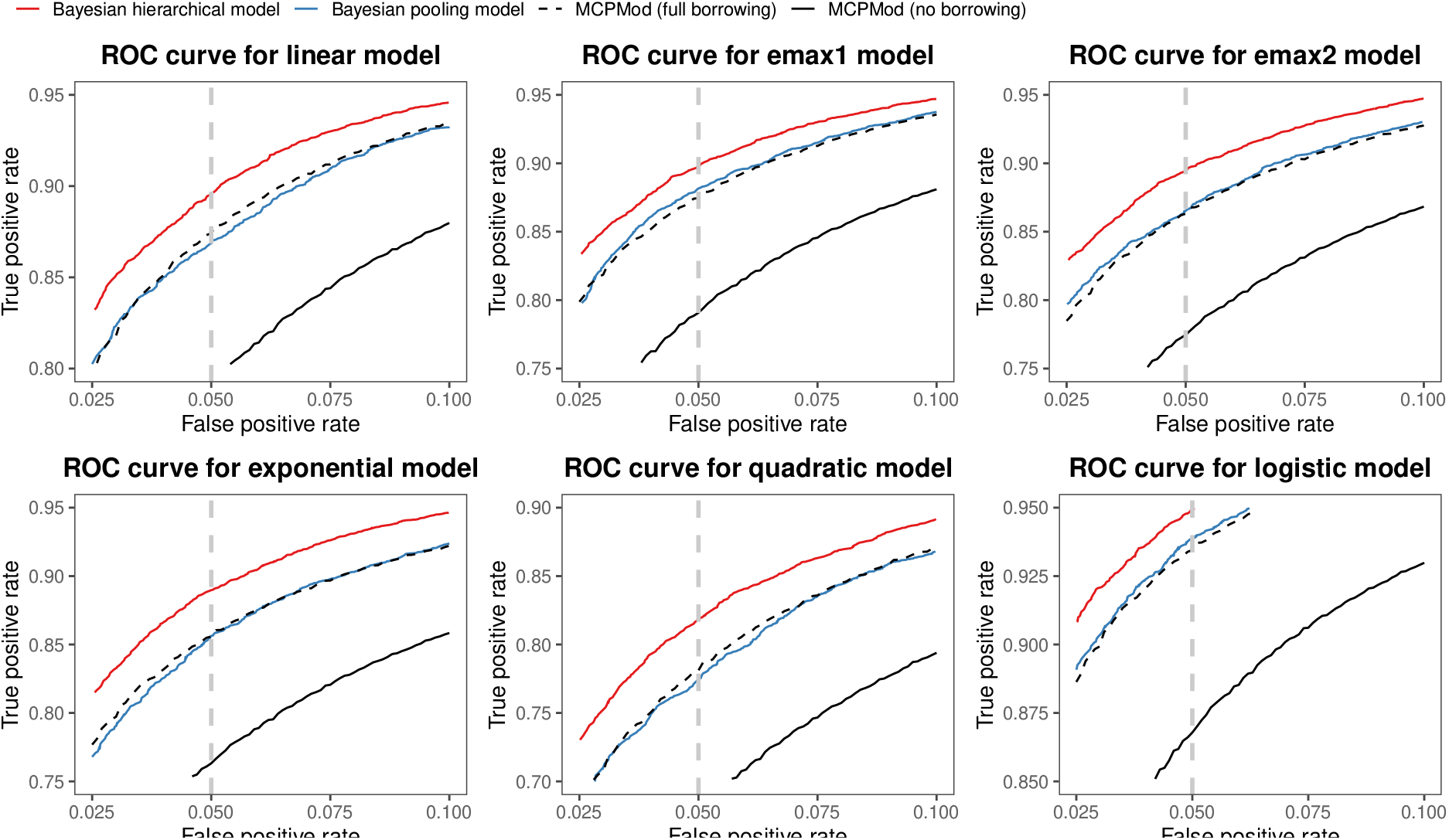
The ROC curves of MCPMod, Bayesian pooling model (BPM) and Bayesian hierarchical model (BHM) across six candidate models for scenario 1A.

**Figure 5:**
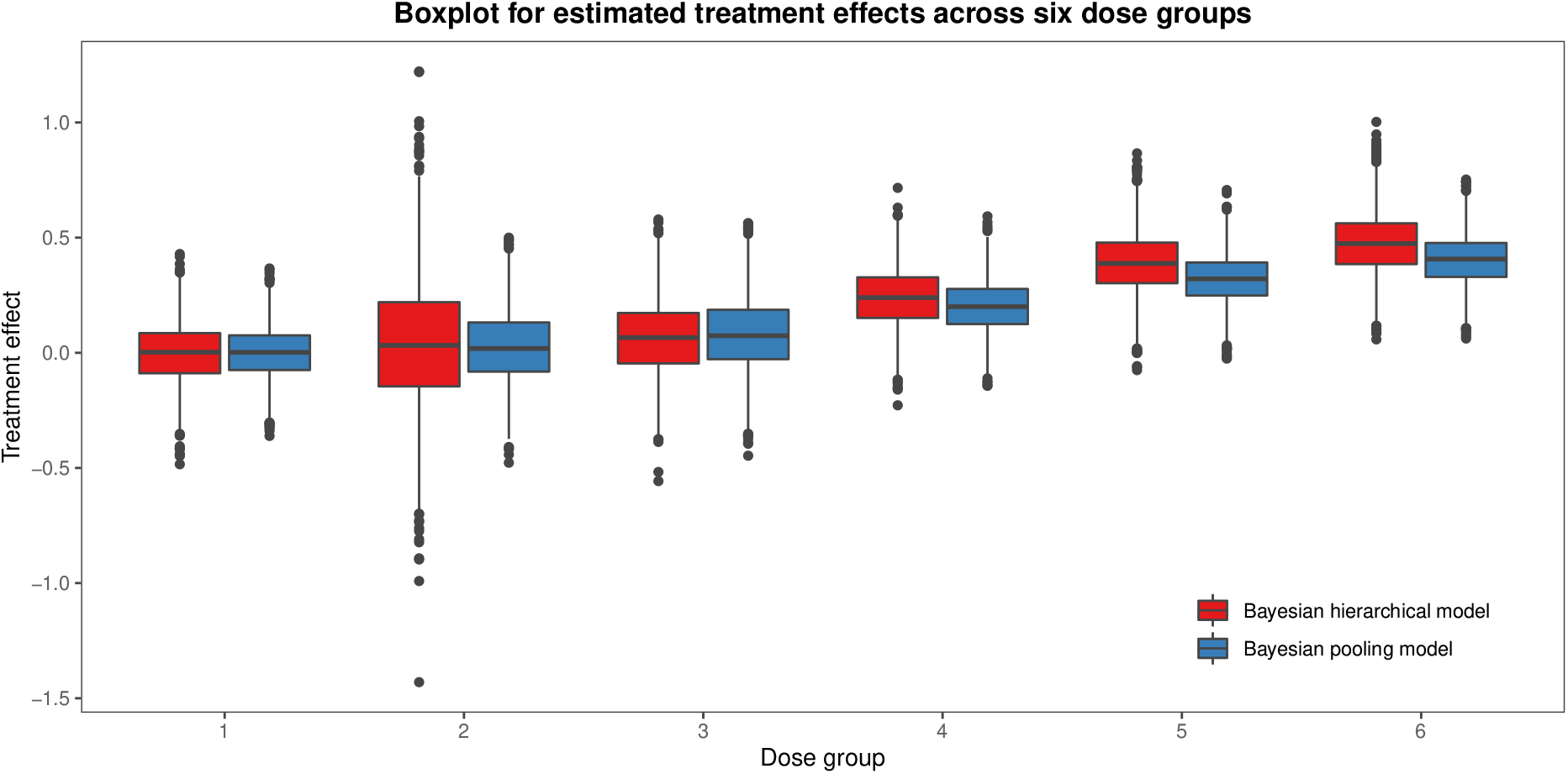
Boxplot of estimated treatment effects across different doses of Bayesian hierarchical model (BHM) and Bayesian pooling model (BPM) under scenario 2A.

Under scenario 2C, we consider two decision rules to detect a significant PoC. The first one is using all of dose groups in current and historical trials (decision rule I) and the second one is using dose groups in the current trial only (decision rule II). The third row of Figure 3 compares the performance of BPM and BHM using these two decision rules. It shows that the ROC curve of BHM is above that of BPM with either decision rule. This is not surprising since BHM produces much more accurate estimates than BPM under scenario 2C, as shown in Table 3. We observe that BHM has better performance when using decision rule II as the dose found only in the historical trial is ignored. This indicates that even though we use a re-estimated contrast vector, the performance is also influenced by the non-overlapping doses. On the other hand, the ROC curve of BPM is higher with decision rule I than decision rule II. Therefore, when there exist non-overlapping doses between the historical and current trials, we recommend using decision rule II when the data is pooled.

## 4. Discussion

In this paper, we proposed a Bayesian hierarchical model for dose finding trials that incorporates information from historical trials. The model can take into account both prognostic and predictive between-trial heterogeneities. We detailed how to set up a Bayesian multiple comparison procedure, how to choose contrast vectors, and how to check for a non-flat dose response given a desired type I error *α*. We evaluated the performance of our model and illustrated the utility of our approach through three simulation studies, in comparison with the Bayesian pooling model, where data from both trials are pooled.

The main advantage of Bayesian hierarchical model lies in scenarios where there is a measurable difference between the behaviour of the historical and the new trials. Compared to the Bayesian pooling model, the Bayesian hierarchical model produces more accurate estimates of the treatment effects in our simulation studies, even when the effect sizes are different in the two trials. We used heavy-tailed hyperpriors for prognostic heterogeneity and truncated normal priors for predictive heterogeneity so that the results were not sensitive to the prior distribution. Even with an inappropriate prior distribution, power usually does not differ too much, hence power values should not be strongly affected by prior-data conflicts. We provide more details on effects of prior-data conflicts in Appendix E.

One key limitation of our approach is that we assumed a fixed effect ratio across different doses, which implies that a fixed relationship between the current and historical trials regardless of dose. In practice, however, this may not hold. This assumption can be relaxed and our approach can be extended to handle this situation, but extended model will have a larger number of unknown parameters. Our method under the assumption of a fixed effect ratio across different doses may be a good trade-off between model complexity and generality. Furthermore, in this work, we only consider the case of borrowing information from one historical dose finding trial. Our method can be easily extended to cases where information from multiple historical trials or parallel trials can be borrowed.

## Data Availability

All data produced in the present work are contained in the manuscript

## A. Metropolis-Hasting algorithm

For each time step *t*, our algorithm consists of the following steps:

Step 1: Generate a proposal sample ***θ***^cand^ = (***μ***^cand^, *a*^cand^, *r*^cand^, *τ*^cand^) from the proposal distribution *q* (***θ***^*t*^ | ***θ***^*t*−1^);
Step 2: Compute the acceptance probability via the acceptance function

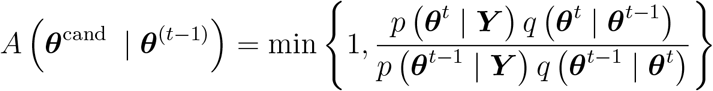

based on the proposal distribution and the joint density function (4).
Step 3: Accept the candidate sample with probability *A*, i.e. take ***θ***^*t*−1^ = ***θ***^cand^. Otherwise, reject the candidate sample and take ***θ***^*t*−1^ = ***θ***^*t*−1^.

These steps are repeated until a sufficient number of posterior samples have been generated.

We run 500, 000 MCMC iterations for each replicate. We discard the first 5, 000 iterations as burn-in and then thin the remaining output by keeping every tenth observation.

## B. Steps for estimating power

The steps for estimating power of our algorithm for fixed values of *a, r* and *α* are as follows:

Step 1: repeat the following steps 10000 times:

1a. Generate ***Y*** ^(*c*)^ and ***Y*** ^(*h*)^ assuming model *m* to be flat, using (1).
1b. Repeat steps of Metropolis-Hastings (MH) algorithm described above until a sufficient number of samples of the parameters ***μ*** have been obtained.
1c. Calculate the probability 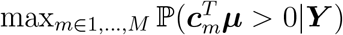.

Step 2: use points with a spacing of 0.0001 from 0 to 1 as thresholds to estimate type I errors (i.e., 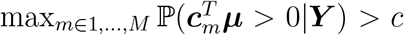, for *c* ∈ [0, 1]) and choose a specific threshold *α* such that the desired type I error 5% is achieved.

Step 3: repeat the following steps 10000 times:

3a. Generate ***Y*** ^(*c*)^ and ***Y*** ^(*h*)^ assuming model *m* to be true, using (1).
3b. Repeat steps of Metropolis-Hastings (MH) algorithm described above until a sufficient number of samples of the parameters ***μ*** have been obtained.
3c. Calculate the probability 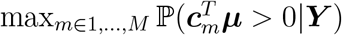.

Step 4: use the obtained threshold *α* in Step 2 and count how many times the test decision is significant (i.e., 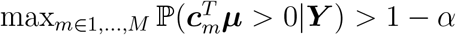, this proportion of simulations for which the null hypothesis is rejected is the estimate of the power of the test.

## C. Figures

### C.1. Figure for scenario 1

### C.2. Figure for scenario 2

## D. Scenario 3: a phase II PoC trial

Scenario 3 is a phase II PoC trial, where two high doses are compared to a placebo dose (i.e., 𝒟_*h*_ = {0, 0.8, 1}). We run our method with same settings as in Section 3.

Table 4 displays power across six candidate models when type I error is controlled at 5%. We generate the response data under six dose-response models in Figure 1 as the true underlying dose-response model without prognostic heterogeneity (*r* = 0). We apply three methods, including MCPMod (no borrowing), BPM (pooling) and BHM (dynamic borrowing), to the dataset under scenario 3A. Both BPM and BHM result in a higher power than the no borrowing model for all of six candidate models. Under 5 of the 6 dose-response models, BHM outperforms BPM. The only exception is the quadratic model. The ROC curves of six candidate models are shown in Figure 6.

**Table 4:**
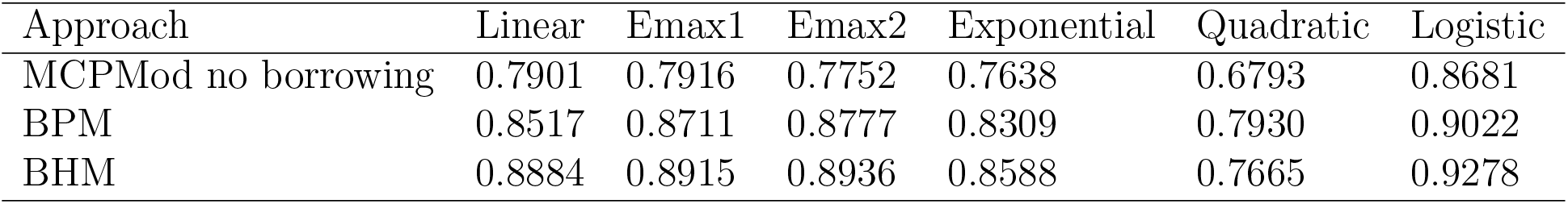
Power values for MCPMod (no borrowing), Bayesian pooling model (BPM) and Bayesian hierarchical model (BHM) for the candidate model set when controlling type I error at 5%.

**Figure 6:**
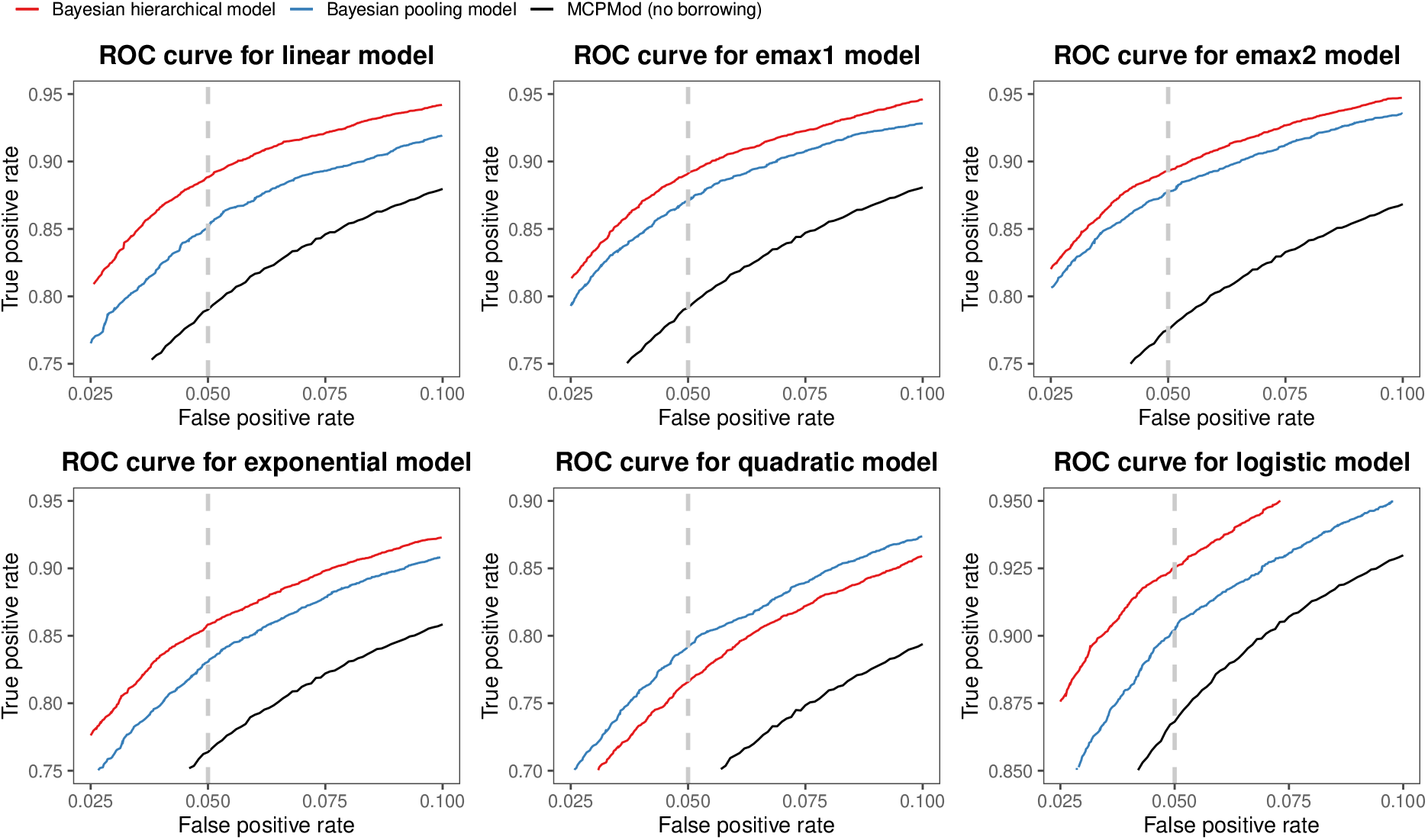
The ROC curves of MCPMod (no borrowing), Bayesian pooling model (BPM) and Bayesian hierarchical model (BHM) across six candidate models for scenario 3A.

Figure 7 shows the ROC curves of MCPMod, BPM and BHM with four levels of prognostic heterogeneity for scenario 3. Results and finding for BHM under scenario 3 are similar to scenarios 1 and 2. However, the behaviour of BPM is different from scenarios 1 and 2. Under scenarios 3A and 3C, as the level of prognostic heterogeneity increases, the ROC curve of BPM lower significantly indicating that it is more sensitive to the heterogeneity of prognostic effects. Table 5 shows the estimated treatment effects of non-overlapping dose groups. It shows BHM produces a much smaller bias in the treatment effect estimates across nearly all scenarios and level of prognostic heterogeneity only has little influence on the treatment effect estimates. On the other hand, BPM produces a large bias in the treatment effect estimates of non-overlapping dose groups and as the level of prognostic heterogeneity increases, there is an obvious increase in bias under all scenarios. There is a large bias in the estimated treatment effects of non-overlapping dose groups using BPM. These two non-overlapping dose groups are in the current trial only, which means that when the effect size of the current trial is larger than that in the historical trial, BPM overestimates the treatment effects of these two dose groups. In contrast, when the effect size of the current trial is smaller than that in the historical trial, BPM underestimates the treatment effect of these two dose groups. Under scenario 3B, Figure 7 shows that the gap between ROC curves of BPM and BHM increase significantly as the prognostic heterogeneity increases. The power values at 5% type I error for BPM increases gradually from 0.8682 for *r* = 0 to 0.9616 for *r* = 0.3. The optimal contrast coefficients of these two dose groups for the linear dose-response curve are negative. But the estimates of *μ*_2_ and *μ*_3_ under the alternative hypothesis have the wrong sign. This leads to an increase in the power of BPM.

**Table 5:** Estimated treatment effect of two non-overlapping dose groups under three simulation scenarios with four different levels of prognostic heterogeneity. BHM = Bayesian hierarchical model; BPM = Bayesian pooling model.

| Scenario | Model | $\vert r\vert = 0$ | $\vert r\vert = 0.1$ | $\vert r\vert = 0.2$ | $\vert r\vert = 0.3$ |
| --- | --- | --- | --- | --- | --- |
| Scenario 3A |  |  |  |  |  |
| Null hypothesis | | $r = 0$ | $r = 0.1$ | $r = 0.2$ | $r = 0.3$ |
| 0.15 ($\mu_2 = 0$) | BHM | 0.0014 | 0.0150 | 0.0219 | 0.0228 |
|  | BPM | 0.0023 | 0.1000 | 0.1996 | 0.2994 |
| 0.50 ($\mu_3 = 0$) | BHM | 0.0015 | 0.0182 | 0.0177 | 0.0252 |
|  | BPM | 0.0006 | 0.1033 | 0.1988 | 0.3024 |
| Alternative hypothesis | | $r = 0$ | $r = 0.1$ | $r = 0.2$ | $r = 0.3$ |
| 0.15 ($\mu_2 = 0.075$) | BHM | 0.0737 | 0.0987 | 0.1098 | 0.1119 |
|  | BPM | 0.0713 | 0.1738 | 0.2738 | 0.3702 |
| 0.50 ($\mu_3 = 0.250$) | BHM | 0.2541 | 0.2730 | 0.2879 | 0.2873 |
|  | BPM | 0.2504 | 0.3525 | 0.4484 | 0.5511 |
| Scenario 3B |  |  |  |  |  |
| Null hypothesis | | $r = 0$ | $r = -0.1$ | $r = -0.2$ | $r = -0.3$ |
| 0.15 ($\mu_2 = 0$) | BHM | 0.0014 | 0.0150 | 0.0219 | 0.0228 |
|  | BPM | 0.0023 | 0.1000 | -0.1996 | -0.2994 |
| 0.50 ($\mu_3 = 0$) | BHM | 0.0015 | 0.0182 | 0.0177 | 0.0252 |
|  | BPM | 0.0006 | -0.1033 | -0.1988 | -0.3024 |
| Alternative hypothesis | | $r = 0$ | $r = -0.1$ | $r = -0.2$ | $r = -0.3$ |
| 0.15 ($\mu_2 = 0.045$) | BHM | 0.0960 | 0.0829 | 0.0812 | 0.0791 |
|  | BPM | 0.0437 | -0.0578 | -0.1510 | -0.2551 |
| 0.50 ($\mu_3 = 0.150$) | BHM | 0.1999 | 0.1937 | 0.1934 | 0.1902 |
|  | BPM | 0.1526 | 0.0513 | -0.0473 | -0.1554 |
| Scenario 3C |  |  |  |  |  |
| Null hypothesis | | $r = 0$ | $r = 0.1$ | $r = 0.2$ | $r = 0.3$ |
| 0.15 ($\mu_2 = 0$) | BHM | -0.0012 | 0.0290 | 0.0576 | 0.0796 |
|  | BPM | 0.0045 | 0.0991 | 0.2010 | 0.3010 |
| 0.50 ($\mu_3 = 0$) | BHM | 0.0010 | 0.0307 | 0.0526 | 0.0757 |
|  | BPM | 0.0013 | 0.0964 | 0.1979 | 0.2960 |
| Alternative hypothesis | | $r = 0$ | $r = 0.1$ | $r = 0.2$ | $r = 0.3$ |
| 0.15 ($\mu_2 = 0.075$) | BHM | 0.0737 | 0.0987 | 0.1098 | 0.1119 |
|  | BPM | 0.0713 | 0.1738 | 0.2738 | 0.3702 |
| 0.50 ($\mu_3 = 0.250$) | BHM | 0.2541 | 0.2730 | 0.2879 | 0.2873 |
|  | BPM | 0.2504 | 0.3525 | 0.4484 | 0.5511 |

**Figure 7:**
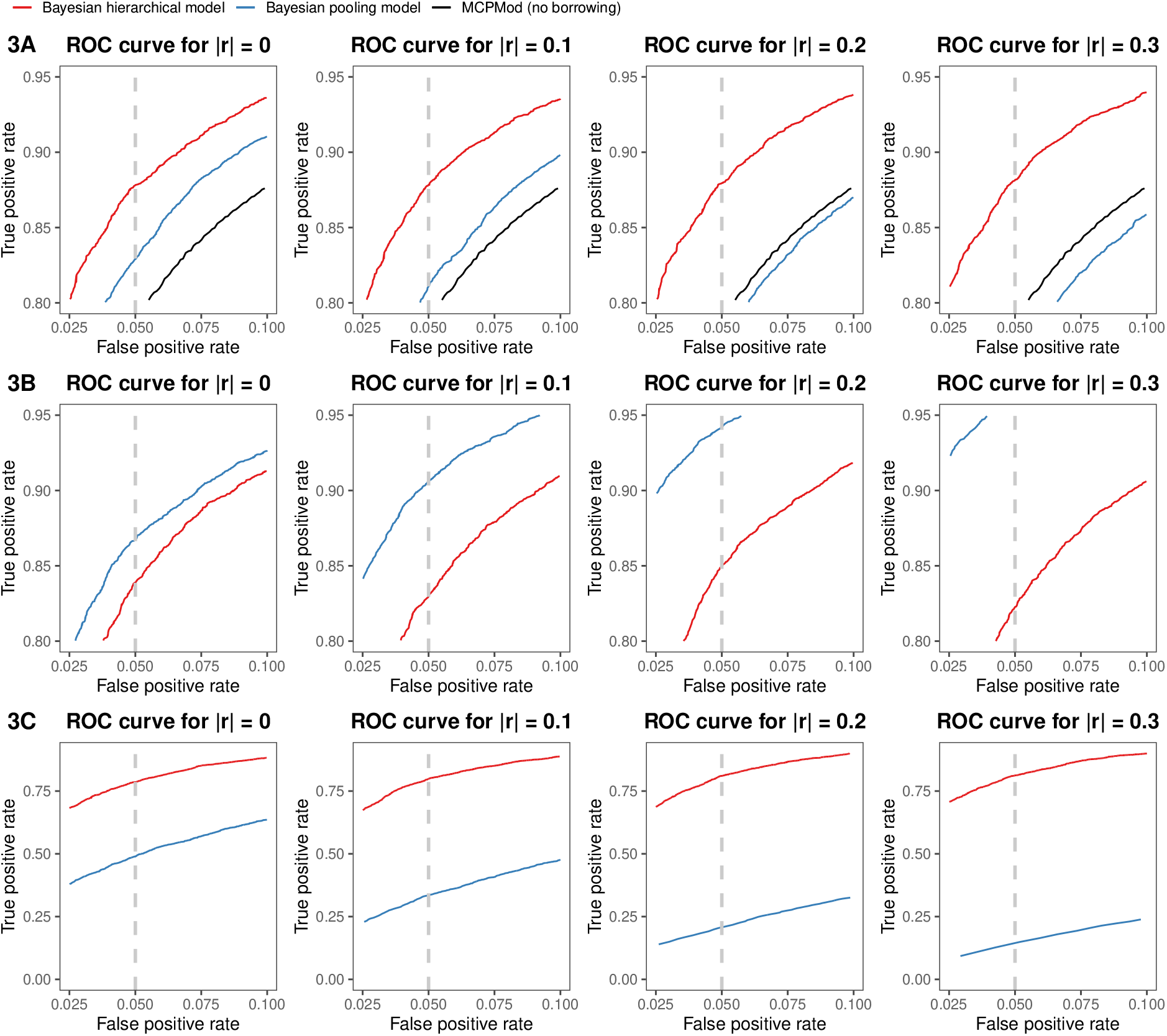
The ROC curves of MCPMod, Bayesian pooling model (BPM) and Bayesian hierarchical model (BHM) with four different levels of prognostic heterogeneity for scenario 3.

In summary, our simulation studies show that BHM can estimate treatment effects of each dose group more accurately than BPM when there exists heterogeneity of prognostic effects, especially for non-overlapping dose groups, resulting in higher statistical power for fixed type I error.

## E. Prior-data conflicts

For the historical trial, there is usually a reasonable amount of information from internal or external company databases, literature and expert opinions that can be used to formulate an appropriate prior of prognostic and predictive heterogeneities. There may exist a mismatch observed between the prior and the data. For this purpose this section will investigate the impact of prior-data conflicts on the performance of BHM under scenarios 1A, 2A and 3A.

We will first discuss prior-data conflict of heterogeneity of prognostic effects. We assume that information on prognostic factors from the current and historical trials can be constructed as informative hyperpriors. In the following simulations, we fix the standard deviation of prior of the effect size *a* at *η* = 0.4 and generate data from two extreme values of *r* = 0 and *r* = 0.5. For the heterogeneity of prognostic effects, we consider five half-normal prior distributions with scales 0.0625, 0.125, 0.25, 0.5 and 1.0 for the prognostic between-study standard deviation *τ*. These specifications include up to small, moderate, substantial, large and very large heterogeneity, respectively.

Table 6 summarises effect size estimates using half-normal prior distribution with five different scales. Generally speaking, when the scale of half-normal distribution is approximately equal to the value of prognostic effect *r*, incorporating historical data on the treatment effect leads to little bias in the estimate. For example, in the case of *r* = 0, effect size estimates using scale 0.0625 were more accurate than other scales. This is because more accurate hyperparameters for the hyper priors are used resulting in more accurate effect size estimates for the current trial. The scale of the half-normal distribution exceeding the value of prognostic effect will result in overestimates the effects size with increasing bias as the level of prognostic heterogeneity increases.

**Table 6:** Estimated treatment effect size using different scales of half-normal prior distribution for three scenarios. The true effect size is 0.5 (Δ_*c*_ = 0.5).

| Scale of half-normal<br>prior distribution | 1 | 0.5 | 0.25 | 0.125 | 0.0625 |
| --- | --- | --- | --- | --- | --- |
| $r = 0$ | | | | | |
| Scenario 1A | 0.4821 | 0.4854 | 0.4815 | 0.4991 | 0.5013 |
| Scenario 2A | 0.4912 | 0.4913 | 0.4997 | 0.4997 | 0.5009 |
| Scenario 3A | 0.4638 | 0.4645 | 0.4766 | 0.4868 | 0.4946 |
| $r = 0.5$ | | | | | |
| Scenario 1A | 0.4912 | 0.4913 | 0.5067 | 0.5152 | 0.5273 |
| Scenario 2A | 0.4952 | 0.5017 | 0.5118 | 0.5203 | 0.5287 |
| Scenario 3A | 0.4920 | 0.5052 | 0.5190 | 0.5249 | 0.5291 |

Table 7 shows power values for the linear model of the candidate model set using half-normal distribution with five levels of prognostic heterogeneity in the hyperprior for the between-trial standard deviation *τ*. When there is no prior-data conflict, the effect size can be estimated more accurately using the appropriate prior distribution, resulting in higher power. In contrast, as the discrepancy between the true prognostic effect of the data and the prior increases, so does the probability of prior-data conflict, which can lead to increases in bias and losses in power. However, power values of half-normal prior distribution with different scales differ slightly, the difference between the maximum power and the minimum power is approximately only 0.1. Using heavy-tailed distributions as hyperpriors has the advantage of ensuring some degree of robustness against prior misspecification, reducing the effects of prior-data conflicts. Even if the historical trial is similar to the current study in their specification, we believe that prior-data conflict may still occur because of additional unanticipated factors. In such situations we recommend using weakly informative half-normal priors *HN* (0.5) that captures heterogeneity values typically seen in meta-analyses of heterogeneous studies and will therefore be a sensible choice in many applications.

**Table 7:** Power values at 5% type I error using different scales of half-normal prior distribution for three scenarios.

| Scale of half-normal<br>prior distribution | 1 | 0.5 | 0.25 | 0.125 | 0.0625 |
| --- | --- | --- | --- | --- | --- |
| $r = 0$ | | | | | |
| Scenario 1A | 0.8891 | 0.8917 | 0.8905 | 0.8935 | 0.8943 |
| Scenario 2A | 0.8853 | 0.8879 | 0.8851 | 0.8859 | 0.8893 |
| Scenario 3A | 0.8689 | 0.8769 | 0.8806 | 0.8831 | 0.8864 |
| $r = 0.5$ | | | | | |
| Scenario 1A | 0.8816 | 0.8922 | 0.8899 | 0.8892 | 0.8835 |
| Scenario 2A | 0.8831 | 0.8905 | 0.8831 | 0.8827 | 0.8773 |
| Scenario 3A | 0.8701 | 0.8769 | 0.8699 | 0.8667 | 0.8614 |

Next, we consider the prior-data conflict for the heterogeneity of predictive effects. As discussed in Section 2.5, the choice of the value of the standard deviation *η* is critical. In these simulations, we use *HN* (0.5) as the prior of prognostic between-heterogeneity standard deviation *τ* and generate data from *r* = 0. For the heterogeneity of treatment effects (i.e., the effects ratio *a*), we consider truncated normal distribution with the same mean 1 and seven values of standard deviation *η*.

Table 9 contains power values for the linear model of the candidate set using seven standard deviations for the prior distributions. Considering the seven prior distributions an impact of the prior-data conflict can be observed. From Table 8 we can see that estimated effect size using BHM is similar to BPM if a lower standard deviation (*η* = 0.1) is used for the prior of *a*. This prior distribution (*TN* (1, 0.1^2^)) represents a situation where we would intuitively consider there is prior-data conflict, because under this prior, the true value of the effect size (*a* = 0.6) cannot be covered, leading to lower power values. In the case of a too high standard deviation (e.g., *η* = 2) there is no longer a benefit through the use of BHM. This is because weakly informative prior distributions should have little influence on the posterior distribution and therefore on the Bayesian inference. As shown in Table 8, we see that values of 0.3, 0.4, 0.5 and 1 have higher power than BPM in all scenarios and the power is higher when the value of *η* is set to 0.4. As mentioned in Section 2.5, the value of *η* can be estimated roughly using the empirical rule and this rule states that 68% of the distribution will occur within one standard deviation. Under scenario A, the true value of effect ratio *a* is equal to 0.6 and a prior distribution *TN* (1, 0.4^2^) has 68% of its observations within one standard deviation of the mean 1 (i.e., 1 *± η*), which means the probability that a variable is within a range [0.6, 1.4] in this normal distribution is 68%. Since this prior distribution is symmetric the standard deviation *η* = 0.4 is also an optimal choice for *a* = 1.4. We recommend using this approach to determine the value of standard deviation *η*.

**Table 8:**
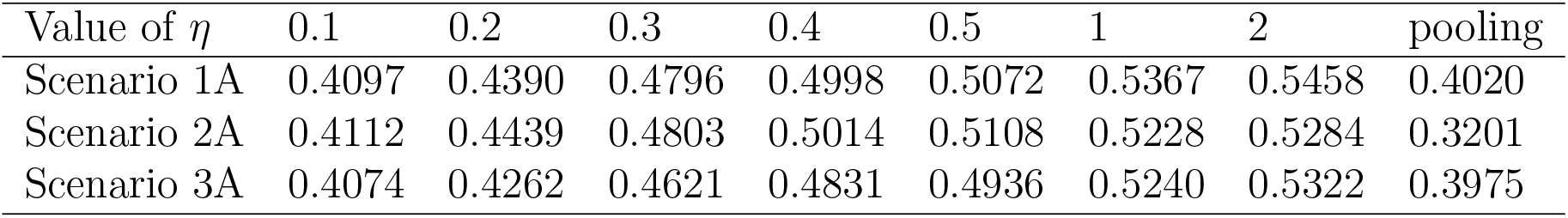
Estimated treatment effect size using different standard deviations of truncated normal prior distribution for three scenarios. The true effect size is 0.5 (Δ_*c*_ = 0.5).

**Table 9:** Power values at 5% type I error using different standard deviations of truncated normal prior distribution for three scenarios.

| Value of $\eta$ | 0.1 | 0.2 | 0.3 | 0.4 | 0.5 | 1 | 2 | pooling |
| --- | --- | --- | --- | --- | --- | --- | --- | --- |
| Scenario 1A | 0.8732 | 0.8845 | 0.8895 | 0.8936 | 0.8891 | 0.8873 | 0.8849 | 0.8753 |
| Scenario 2A | 0.8803 | 0.8829 | 0.8894 | 0.8916 | 0.8897 | 0.8841 | 0.8789 | 0.8828 |
| Scenario 3A | 0.8492 | 0.8605 | 0.8672 | 0.8832 | 0.8762 | 0.8705 | 0.8694 | 0.8461 |

In the case of no reliable information for the effect ratio, the prior distribution needs to be suitably vague so that it includes the unexpected, e.g. *η* = 0.5 and *η* = 1. There exists several appropriate choices of the standard deviation. Even though the inappropriate choice of the prior distribution is selected, the power usually do not differ too much that means power values should not be strongly affected by prior-data conflicts. This is due to weakly informative truncated normal prior distributions used in our model. This truncated normal prior distribution with a truncation range [1/3, 3] provides better robustness than a normal distribution. A value of the effect ratio *a* exceeds this range corresponds to extremely large heterogeneity of treatment effects, and would essentially lead to no borrowing from the historical trial.

Therefore, heavy-tailed hyperpriors for prognostic heterogeneity and truncated normal priors for predictive heterogeneity imply a degree of robustness against prior-data conflicts which means the results are not sensitive to the prior specification. Even though the inappropriate choice of the prior distribution is selected, the power usually do not differ too much that means power values should not be strongly affected by prior-data conflicts.

## References

Berry, D. A. (2006). Bayesian clinical trials. Nature Reviews Drug Discovery, 5, 27–36.

Bradley, A. P. (1997). The use of the area under the ROC curve in the evaluation of machine learning algorithms. Pattern Recognition Letters, 30, 1145–1159.

Bretz, F., Pinheiro, J. C., & Branson, M. (2005). Combining multiple comparisons and modeling techniques in dose-response studies. Biometrics, 61, 738–748.

European Medicines Agency (EMA) (2014). Qualification Opinion of MCP-Mod as an efficient statistical methodology for model-based design and analysis of Phase II dose finding studies under model uncertainty. https://www.ema.europa.eu/en/documents/regulatory-procedural-guideline/qualification-opinion-mcp-mod-efficient-statistical-methodology-model-based-designen.pdf.

Fawcett, T. (2006). An introduction to ROC analysis. Pattern Recognition Letters, 27, 861–874.

Fleischer, F., Bossert, S., Deng, Q., Loley, C., & Gierse, J. (2022). Bayesian MCPMod. Pharmaceutical Statistics,. doi:10.1002/pst.2193.

Friede, T., Röver, C., Wandel, S., & Neuenschwander, B. (2017). Meta-analysis of two studies in the presence of heterogeneity with applications in rare diseases. Biometrical Journal, 59, 658–671.

Gelman, A. (2006). Prior distributions for variance parameters in hierarchical models. Bayesian Analysis, 1, 515–534.

Hobbs, B. P., Sargent, D. J., & Carlin, B. P. (2012). Commensurate priors for incorporating historical information in clinical trials using general and generalized linear models. Bayesian Analysis, 7, 639–674.

Ibrahim, J. G., & Chen, M.-H. (2000). Power prior distributions for regression models. Statistical Science, 15, 46–60.

Neuenschwander, B., Capkun-Niggli, G., Branson, M., & Spiegelhalter, D. J. (2010). Summarizing historical information on controls in clinical trials. Clinical Trials, 7, 5–18.

Pinheiro, J., Bornkamp, B., Glimm, E., & Bretz, F. (2014). Model-based dose finding under model uncertainty using general parametric models. Statistics in Medicine, 33, 1646–1661.

Polson, N. G., & Scott, J. G. (2012). On the half-Cauchy prior for a global scale parameter. Bayesian Analysis, 7, 887–902.

Rover, C., Bender, R., Dias, S., Schmid, C. H., Schmidli, H. et al. (2021). On weakly informative prior distributions for the heterogeneity parameter in Bayesian random-effects meta-analysis. Research Synthesis Methods, 12, 448–474.

Schmidli, H., Gsteiger, S., Roychoudhury, S., O’Hagan, A., Spiegelhalter, D. et al. (2014). Robust meta-analytic-predictive priors in clinical trials with historical control information. Biometrics, 70, 1023–1032.

Spiegelhalter, D. J., Abrams, K. R., & Myles, J. P. (2004). Bayesian Approaches to Clinical Trials and Health-care Evaluation. Chichester: John Wiley & Sons.

USA Food and Drug Administration (FDA) (2016). Statistical review and evaluation qualification of statistical approach: MCP-Mod. https://www.fda.gov/downloads/Drugs/DevelopmentApprovalProcess/UCM508701.pdf.

